# Prognostic accuracy of emergency department triage tools for children with suspected COVID-19: The PRIEST observational cohort study

**DOI:** 10.1101/2020.09.01.20185793

**Authors:** The PRIEST Research Group, Katie Biggs, Ben Thomas, Steve Goodacre, Ellen Lee, Laura Sutton, Matthew Bursnall, Amanda Loban, Simon Waterhouse, Richard Simmonds, Carl Marincowitz, Jose Schutter, Sarah Connelly, Elena Sheldon, Jamie Hall, Emma Young, Andrew Bentley, Kirsty Challen, Chris Fitzimmons, Tim Harris, Fiona Lecky, Andrew Lee, Ian Maconochie, Darren Walter

**Affiliations:** University of Sheffield, UK; Manchester University NHS Foundation Trust, Wythenshawe Hospital, UK; Lancashire Teaching Hospitals NHS Foundation Trust, UK; Sheffield Children’s NHS Foundation Trust, UK; Barts Health NHS Trust, UK; Imperial College Healthcare NHS Trust, UK

## Abstract

**Objectives:** Emergency department clinicians can use triage tools to predict adverse outcome and support management decisions for children presenting with suspected COVID-19. We aimed to estimate the accuracy of triage tools for predicting severe illness in children presenting to the emergency department (ED) with suspected COVID-19 infection.

**Methods:** We undertook a mixed prospective and retrospective observational cohort study in 44 EDs across the United Kingdom (UK). We collected data from children attending with suspected COVID-19 between 26 March 2020 and 28 May 2020, and used presenting data to determine the results of assessment using the WHO algorithm, swine flu hospital pathway for children (SFHPC), Paediatric Observation Priority Score (POPS) and Children’s Observation and Severity Tool (COAST). We recorded 30-day outcome data (death or receipt of respiratory, cardiovascular or renal support) to determine prognostic accuracy for adverse outcome.

**Results:** We collected data from 1530 children, including 26 (1.7%) with an adverse outcome. C-statistics were 0.80 (95% confidence interval 0.73-0.87) for the WHO algorithm, 0.80 (0.71-0.90) for POPS, 0.76 (0.67-0.85) for COAST, and 0.71 (0.59-0.82) for SFHPC. Using pre-specified thresholds, the WHO algorithm had the highest sensitivity (0.85) and lowest specificity (0.75), but POPS and COAST could optimise sensitivity (0.96 and 0.92 respectively) at the expense of specificity (0.25 and 0.38 respectively) by using a threshold of any score above zero instead of the pre-specified threshold.

**Conclusion:** Existing triage tools have good but not excellent prediction for adverse outcome in children with suspected COVID-19. POPS and COAST could achieve an appropriate balance of sensitivity and specificity for supporting decisions to discharge home by considering any score above zero to be positive.

**Registration:** ISRCTN registry, ISRCTN28342533, http://www.isrctn.com/ISRCTN28342533

## Introduction

COVID-19 causes mild illness in children, compared to adults, with less than 1% of those aged under 20 needing hospital admission, and case fatality rates of 0.0026% in those aged 0-9 years and 0.0148% in those aged 10-19. [1] This still creates a challenge for clinicians assessing children attending the emergency department (ED) with suspected COVID-19, who need to identify and admit the rare cases with severe illness, while allowing safe discharge for the majority with mild illness [2,3].

Triage tools can support decision-making for children presenting to the ED with acute illness. They combine information from clinical assessment in a structured manner to predict the risk of adverse outcome. Paediatric early warning scores are a form of triage tool that use clinical information to generate a score, with a higher score indicating a higher risk of adverse outcome. A number of paediatric early warning scores have been developed and evaluated in general paediatric populations, but with insufficient evidence to recommend one over another [4]. We selected two early warning scores for evaluation as triage tools for suspected COVID-19:

1. Roland et al developed the Paediatric Observation Priority Score (POPS) [5,6] to aid detection of serious illness in paediatric EDs. It consists of eight domains (oxygen saturations, level of alertness, extent of breathing difficulty, background history, nurse gut feeling, heart rate, respiratory rate and temperature) each graded zero, one or two to give a total score of 16.
2. The Children’s Observation and Severity Tool (COAST) [7] was developed from an existing paediatric early warning score for use in the ED. It consists of seven domains (doctor/nurse/family concern, heart rate for age, respiratory rate, oxygen saturation, respiratory distress, altered consciousness and pain score) each graded zero or one to give a total score of seven.

Triage tools can also take the form of an algorithm or set of criteria that use clinical information to generate a positive or negative overall assessment. The World Health Organisation (WHO) developed a decision-making algorithm for hospitalisation of children with COVID-18 pneumonia [8]. It recommends hospitalisation if specified criteria are met, based on respiratory rate, oxygen saturation, respiratory distress, respiratory exhaustion, severe dehydration, reduced conscious level or any comorbidities (diabetes, cardiovascular disease, chronic respiratory disease, renal impairment, immunosuppression). The United Kingdom (UK) Department of Health developed the Swine Flu Hospital Pathway for the 2009 H1N1 influenza pandemic [9]. It recommends hospitalisation if specified criteria are met, based on respiratory distress, respiratory rate, oxygen saturation, respiratory exhaustion, dehydration or shock, altered conscious level, or other clinical concern.

### Aims and objectives

We aimed to estimate the accuracy of triage tools for predicting severe illness in children presenting to the ED with suspected COVID-19 infection.

## Methods

We set up the Pandemic Influenza Triage in the Emergency Department (PAINTED) study following the 2009 H1N1 pandemic to develop and evaluate triage tools for suspected pandemic influenza. We changed it to Pandemic Respiratory Infection Emergency System Triage (PRIEST) study in January 2020 to address any pandemic respiratory infection. The study was activated on 20 March 2020 and collected data across 70 EDs throughout the first wave of the pandemic in the United Kingdom (UK).

PRIEST was an observational cohort study of patients presenting to the ED with suspected COVID-19. We collected standardised predictor variables at presentation and then followed patients up to 30 days after presentation. The study did not involve any change to patient care. Hospital staff made admission and discharge decisions according to usual practice, informed by local and national guidance. Initial descriptive analysis [10] showed that suspected COVID-19 presentations differed markedly between adults and children, and the adverse outcome rate was very low in children. We therefore planned to analyse triage tools separately in adults and children, and not to attempt to derive a new tool for children, given the lack of statistical power.

We identified consecutive patients presenting to the ED of participating hospitals with suspected COVID-19 infection. Patients were eligible if they met the clinical diagnostic criteria [11] of fever (≥ 37.8°C) and acute onset of persistent cough (with or without sputum), hoarseness, nasal discharge or congestion, shortness of breath, sore throat, wheezing, sneezing. This was determined on the basis of the assessing clinician recording that the patient had suspected COVID-19 or completing a standardised assessment form designed for suspected pandemic respiratory infection [12]. We did not seek consent to collect data but information about the study was provided in the ED and patients or parents could withdraw their data at their request. Patients with multiple presentations to hospital were only included once, using data from the first presentation identified by research staff.

We planned to evaluate the WHO algorithm, the Swine Flu Hospital Pathway for children (SFHPC), POPS and COAST. The triage tools are described in Appendix 1. We excluded some variables from the scores on the basis that they were subjective or not relevant to suspected COVID-19, and thus unlikely to be recorded routinely or included in assessment of children with suspected COVID-19. The variables were pain score (COAST), gut feeling (POPS), other clinical cause for concern (SFHPC). We therefore evaluated modified versions of these triage tools. For ease of reading, we have not generally prefixed the scores with the word “modified” but reporting of our findings should recognise that we evaluated modified versions of POPS, COAST and the SFHPC.

Data collection was both prospective and retrospective. Participating EDs were provided with a standardised data collection form that included predictor variables used in existing triage methods or considered to be potentially useful predictors of adverse outcome. Participating sites could adapt the form to their local circumstances, including integrating it into electronic or paper clinical records to facilitate prospective data collection, or using it as a template for research staff to retrospectively extract data from clinical records.

Research staff at participating hospitals reviewed patient records at 30 days after initial attendance to identify any adverse outcomes. Patients who died or required respiratory, cardiovascular or renal support were classified as having an adverse outcome. Patients who survived to 30 days without requiring respiratory, cardiovascular or renal support were classified as having no adverse outcome. Respiratory support was defined as any intervention to protect the patient’s airway or assist their ventilation, including non-invasive ventilation or acute administration of continuous positive airway pressure. It did not include supplemental oxygen alone or nebulised bronchodilators. Cardiovascular support was defined as any intervention to maintain organ perfusion, such as inotropic drugs, or invasively monitor cardiovascular status, such as central venous pressure or pulmonary artery pressure monitoring, or arterial blood pressure monitoring. It did not include peripheral intravenous canulation or fluid administration. Renal support was defined as any intervention to assist renal function, such as haemoperfusion, haemodialysis or peritoneal dialysis. It did not include intravenous fluid administration.

The sample size was dependent on the size and severity of the pandemic, but based on a previous study in the 2009 H1N1 influenza pandemic we estimated we would need to collect data from 20,000 patients across 40-50 hospitals to identify 200 with an adverse outcome, including 50 children with adverse outcome. In the event, the number of children with adverse outcome was insufficient to allow derivation of a new triage tool.

For this analysis, we selected all children (defined as age 15 or younger on date of attendance) for whom we could ascertain presence or absence of adverse outcome at 30 days. We performed the Analysis using Stata v16 [13] and in accordance with a prospective statistical analysis plan. We compared baseline characteristics, presenting features and physiology between children with and without adverse outcome. The study statistician retrospectively applied triage tools to the patient data, as outlined in Appendix 1. We plotted a ROC curve for each triage tool and calculated the area under the ROC curve (c-statistic) for discriminating between cases with and without adverse outcome. We calculated sensitivity, specificity, positive predictive value and negative predictive value at key pre-specified decision-making thresholds. We treated missing data as normal or negative when calculating triage tool scores, but excluded cases from analysis if fewer than three of the factors that make up the score were complete.

## Results

The PRIEST study recruited 1530 children patients across 4 paediatric and 40 mixed EDs between 26 March 2020 and 28 May 2020. The study population had a median age of 2 years (interquartile range 0 to 6), 821 (54.3%) were male and 691 (45.7%) female (18 age missing). Ethnicity was 950 (76.6%) White, 106 (8.5%) Asian, 52 (4.2%) Black/African/Caribbean, 81 (6.5%) mixed/multiple ethnic groups for, and 51 (4.1%) other (missing/prefer not to say 290). After ED assessment, 1109 (72.6%) were discharged, 418 (27.4%) admitted (three missing). Testing for respiratory pathogens identified 19 (1.2%) cases of COVID-19, two (0.1%) cases of influenza and 237 (15.5%) other pathogens. Follow-up data were recorded for 1527 (99.8%) children and identified 26 (1.7%) with an adverse outcome, including four deaths, 18 receiving respiratory support, eight receiving cardiovascular support and two receiving renal support. None of the deaths received organ support but some children received multiple organ support. Adverse outcomes occurred on the same day or within a day of ED assessment for 20 (77%) patients.

Table 1 compares the predictor variables between children with and without adverse outcome, along with the results of univariate analysis. Older age, Black ethnicity and presentation with shortness of breath were associated with increased risk of adverse outcome, while presentation with fever was associated with decreased risk. Co-morbidities were uncommon and only asthma and other chronic lung disease were associated with adverse outcome. Heart rate, respiratory rate, temperature and oxygen saturation were analysed in POPS categories to account for age-related variation in normal ranges and non-linear associations with adverse outcome. Abnormal physiology was associated with increased risk of adverse outcome, as were severe respiratory distress, severe dehydration and abnormal central capillary refill.

**Table 1:**
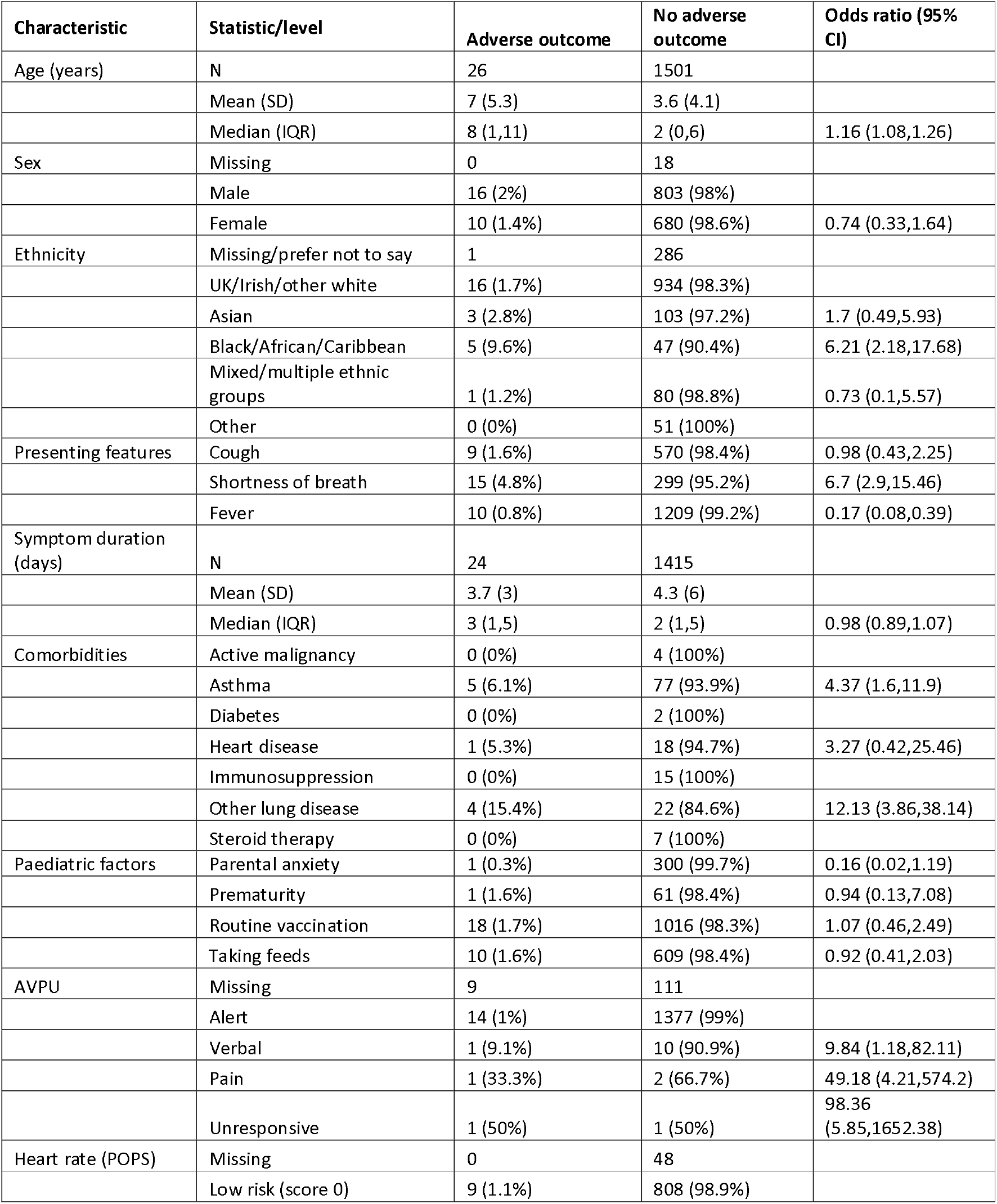
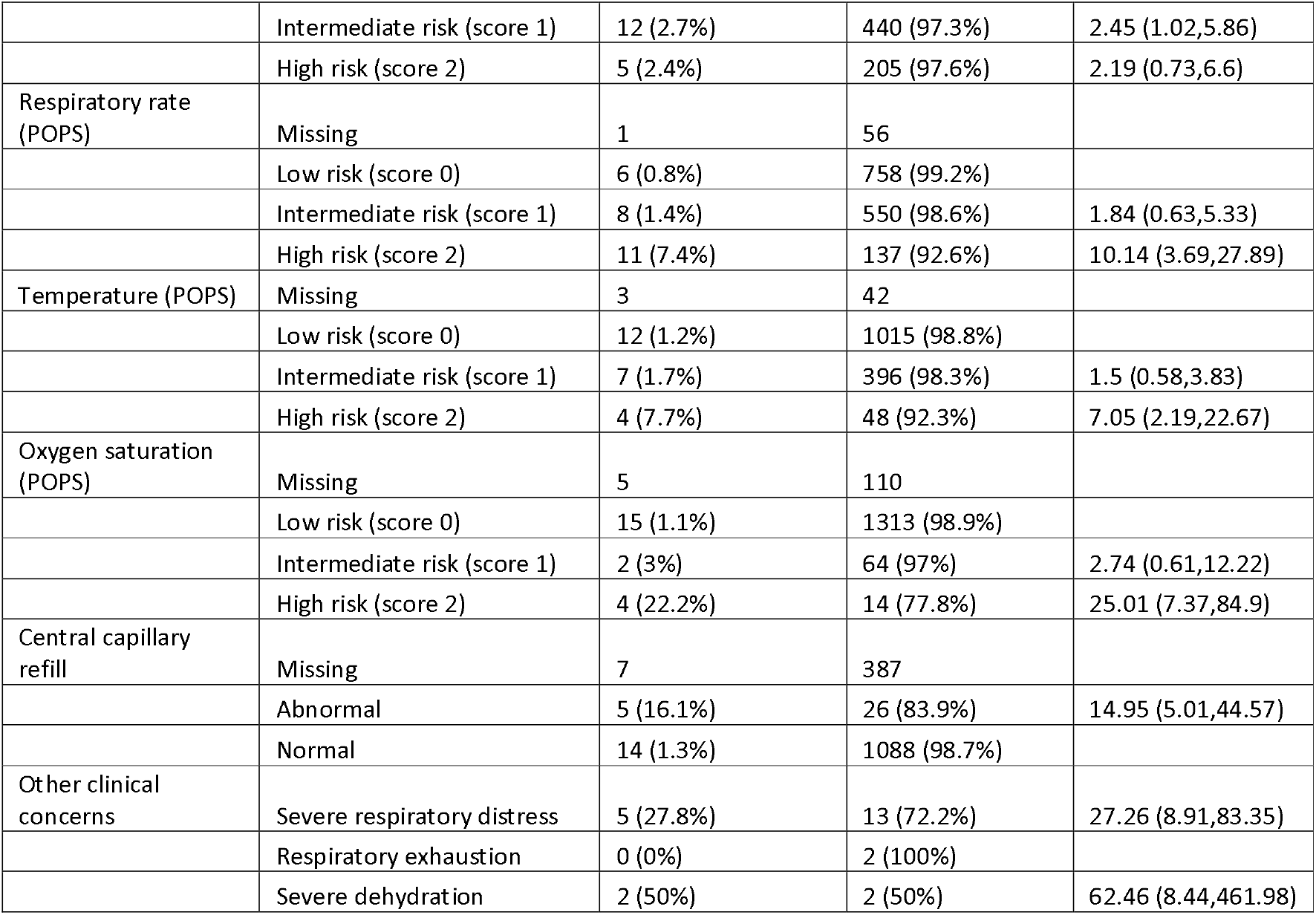
Baseline characteristics, presenting features and physiology of children with (n=26) and without (n=1501)

Figure 1 shows the ROC curves for the triage tools and Table 2 shows c-statistics and the prognostic parameters at the pre-specified thresholds. C-statistics varied from 0.71 (SFHPC) to 0.8 (POPS and WHO algorithm), although confidence intervals overlapped. The triage tools showed varying trade-offs between sensitivity and specificity at the pre-specified thresholds, with the WHO algorithm having highest sensitivity (0.85), while the SFHPC, POPS and COAST showed progressively lower sensitivity and higher specificity. Table 3 shows the sensitivity and specificity of POPS and COAST scores across the range of thresholds. Using any score above zero as the threshold for positivity, POPS would have sensitivity of 0.96 (95% confidence interval [CI] 0.80 to 1.00) and specificity 0.25 (0.23 to 0.27), and COAST would have sensitivity of 0.92 (0.74 to 0.99) and specificity of 0.38 (0.36 to 0.41).

**Figure 1:**
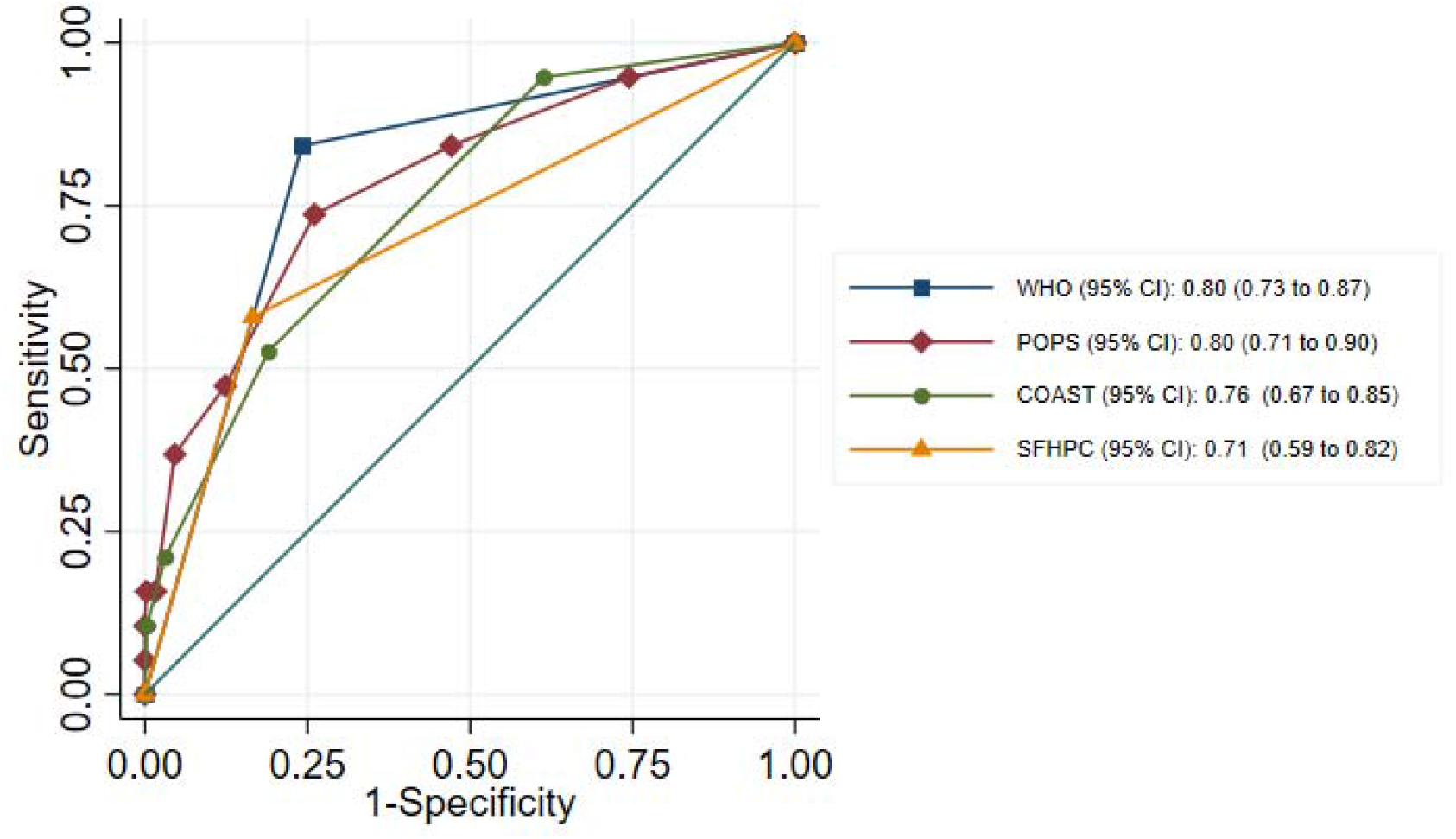
ROC curves with AUC (C-statistic) for predicting adverse outcome

**Table 2:** Predictive accuracy of each triage tool for adverse outcome

| Triage tool | n | C-statistic | Pre-specified threshold | Sensitivity (95% CI) | Specificity (95% CI) | Positive Predictive Value (95% CI) | Negative Predictive Value (95% CI) |
| --- | --- | --- | --- | --- | --- | --- | --- |
| WHO | 1,530 | 0.80<br>(0.73, 0.87) | (≥1) | 0.85<br>(0.65, 0.96) | 0.75<br>(0.73, 0.78) | 0.06<br>(0.04, 0.08) | 1.00<br>(0.99, 1.00) |
| POPS | 1,499 | 0.80 | (≥5) | 0.40 | 0.95 | 0.13 | 0.99 |
|  |  | (0.71, 0.90) |  | (0.21, 0.61) | (0.94, 0.96) | (0.06, 0.22) | (0.98, 0.99) |
| COAST | 1,489 | 0.76 | (≥3) | 0.16 | 0.97 | 0.08 | 0.99 |
|  |  | (0.67, 0.85) |  | (0.05, 0.36) | (0.96, 0.98) | (0.02, 0.19) | (0.98, 0.99) |
| SFHPC | 1,395 | 0.71 | (≥1) | 0.58 | 0.83 | 0.05 | 0.99 |
|  |  | (0.59, 0.82) |  | (0.33, 0.80) | (0.81, 0.85) | (0.02, 0.08) | (0.99, 1.00) |

**Table 3:** Sensitivity and specificity for each observed threshold of COAST and POPS triage tools

| Tool | Threshold | Sensitivity (95% CI) | Specificity (95% CI) |
| --- | --- | --- | --- |
| COAST | ≥1 | 0.92 (0.74 to 0.99) | 0.38 (0.36 to 0.41) |
|  | ≥2 | 0.60 (0.39 to 0.79) | 0.81 (0.79 to 0.83) |
|  | ≥3 | 0.16 (0.05 to 0.36) | 0.97 (0.96 to 0.98) |
|  | ≥4 | 0.08 (0.01 to 0.26) | 1.00 (0.99 to 1.00) |
| POPS | ≥1 | 0.96 (0.80 to 1.00) | 0.25 (0.23 to 0.27) |
|  | ≥2 | 0.84 (0.64 to 0.95) | 0.52 (0.50 to 0.55) |
|  | ≥3 | 0.76 (0.55 to 0.91) | 0.73 (0.71 to 0.75) |
|  | ≥4 | 0.56 (0.35 to 0.76) | 0.88 (0.86 to 0.89) |
|  | ≥5 | 0.40 (0.21 to 0.61) | 0.95 (0.94 to 0.96) |
|  | ≥6 | 0.20 (0.07 to 0.41) | 0.98 (0.97 to 0.99) |
|  | ≥7 | 0.12 (0.03 to 0.31) | 1.00 (0.99 to 1.00) |
|  | ≥8 | 0.08 (0.01 to 0.26) | 1.00 (1.00 to 1.00) |
|  | ≥9 | 0.04 (0.00 to 0.20) | 1.00 (1.00 to 1.00) |

## Discussion

### Summary of main findings

Our findings show that ED triage tools have good but not excellent discriminant value for predicting adverse outcome in children with suspected COVID-19. POPS and the WHO algorithm have higher point estimates for c-statistics than COAST or the SFHPC, but lack of statistical power limits our ability to conclude that one triage tool is better than another. Triage tools need to optimise sensitivity at the expense of specificity if they are used to support decisions to admit to hospital or discharge home, where false negative assessment could lead to delayed treatment. Our findings suggested that the triage tools performed with suboptimal sensitivity at the pre-specified thresholds, but POPS and COAST could optimise sensitivity to potentially acceptable levels by sacrificing specificity and treating any score above zero as positive.

### Previous research

There is limited relevant existing research to compare with our findings. To date, studies have described the characteristics of children with COVID-19 but have not, to our knowledge, evaluated triage tools or early warning scores. Lu et al described 1391 children investigated for suspected COVID-19 in Wuhan, China, of whom 12.3% had infection confirmed, three required intensive care and one died [14]. Bellino et al described 3836 children with COVID-19 in Italy, of whom 13.3% were admitted to hospital, 3.5% admitted to intensive care and 0.1% died [15]. Age below one year and underlying conditions were associated with increased risk of adverse outcome. Gotzinger et al described 582 children with COVID-19 across Europe, of whom 62% were admitted to hospital and 8% to intensive care [16]. Younger age, male sex, pre-existing conditions and presence of symptoms or signs of lower respiratory tract infection prediction intensive care admission on multivariable analysis. Swann et al described 651 children with COVID-19 admitted to UK hospitals, of whom 18% were admitted to critical care and 1% died [17]. Age under one month or between 10 and 14 years, and Black ethnicity predicted critical care admission on multivariable analysis.

We previously evaluated the SFHPC in a cohort consisting mostly of children in the 2009 H1N1 pandemic [18], using the same definition of adverse outcome as this study, and reported a c-statistic of 0.62 (95% CI 0.51 to 0.72), sensitivity of 0.60 (0.23 to 0.88), and specificity of 0.81 (0.73 to 0.87). Roland et al evaluated POPS in 24068 children presenting with any condition to the ED and reported a c-statistic of 0.802 for predicting hospital admission [19]. The majority of patients (68.5%) were POPS zero, of whom only 794 (4.8%) were admitted to hospital and only eleven returned for admission after initial discharge. These findings concur with ours in suggesting that a score of zero versus anything above zero as an appropriate threshold for supporting decisions to admit or discharge. Lillitos et al evaluated COAST in 1921 paediatric ED attendances and reported c-statistics of 0.69 (95% CI 0.65 to 0.73) for hospital admission and 0.75 (0.72 to 0.79) for significant medical illness [7]. They did not report sensitivity and specificity for a threshold of zero versus anything above zero, but reported sensitivity and specificity for hospital admission at our pre-specified threshold (greater or equal to three) as 0.32 (0.25 to 0.38) and 0.93 (0.92 to 0.94) respectively, suggesting a much lower threshold is needed for decisions around hospital admission or discharge.

### Strengths and limitations

We collected data across multiple varied sites throughout the first wave of the pandemic in the UK. This analysis is therefore based on a large and representative sample of children with suspected COVID-19. However, the low rate of adverse outcome (1.7%) meant that our study lacked statistical power to detect associations between predictors and adverse outcome. We were unable to address our original aim of deriving a new triage tool and lacked sufficient adverse outcomes to undertake multivariable analysis. The associations reported in Table 1 are based on univariate analysis and should be considered with caution. The differences between point estimates for c-statistics and sensitivity are relatively imprecise. Another limitation is that we relied on a mixture of prospective and retrospective methods to record predictor variables, which resulted in missing data for some variables and inability to determine whether some predictors were not present or not recorded. This may have resulted in some predictor variables being under-recorded, leading to under-estimation of the performance of the triage tools. It should be noted that we evaluated modified versions of POPS, COAST and the SFHPC that dropped a predictor variable from each tool. Inclusion of these dropped variables could have improved prediction, and improved sensitivity at the expense of specificity. Finally, we may have missed adverse outcomes if patients attended a different hospital after initial hospital discharge. This is arguably less likely in the context of a pandemic, in which movements between regions were curtailed, but cannot be discounted.

### Implications for practice

Children presenting with suspected COVID-19 differ markedly from adults, having much lower rates of COVID-19 positivity, hospital admission and adverse outcome. Triage tools are therefore likely to be used with a low threshold for positivity to optimise sensitivity and support decisions to discharge home. Our findings suggest that children with a POPS or COAST score of zero could be discharged home with a low risk of adverse outcome. Higher thresholds could be used to support inpatient referral decisions, such as critical care review, but should not be used to support decisions to discharge home.

None of the scores showed excellent discriminant value for predicting adverse outcome, so there is potential to develop new scores to improve prediction. However, further research in this direction is likely to be limited by the low rate of adverse outcome. Furthermore, the low rate of COVID-19 positivity and adverse outcome seen in children with suspected COVID-19 suggest that there may be little to be gained from treating children with suspected COVID-19 any differently from those presenting with other febrile illnesses (other than for infection control purposes). Future research may therefore be better focussed on developing and validating general paediatric early warning scores, rather than a specific triage tool for suspected COVID-19.

### Ethical approval

The North West -Haydock Research Ethics Committee gave a favourable opinion on the PAINTED study on 25 June 2012 (reference 12/NW/0303) and on the updated PRIEST study on 23rd March 2020. The Confidentiality Advisory Group of the Health Research Authority granted approval to collect data without patient consent in line with Section 251 of the National Health Service Act 2006.

### Contributor and guarantor information

SG, AB, KC, CF, TH, FL, ALe, IM and DW conceived and designed the study. BT, KB, ALo, SW, RS, JS, SC, ES, JH and EY acquired the data. EL, LS, SG, BT, KB and CM analysed the data. SG, AB, KC, CF, TH, FL, ALe, IM, DW, EL, LS, SG, BT, KB and CM interpreted the data. All authors contributed to drafting the manuscript. Steve Goodacre is the guarantor of the paper. The corresponding author attests that all listed authors meet authorship criteria and that no others meeting the criteria have been omitted.

### Competing interests

All authors have completed the ICMJE uniform disclosure form at www.icmje.org/coi_disclosure.pdf and declare: grant funding to their employing institutions from the National Institute for Health Research; no financial relationships with any organisations that might have an interest in the submitted work in the previous three years; no other relationships or activities that could appear to have influenced the submitted work.

## Data Availability

Anonymised data are available from the corresponding author upon reasonable request (contact details on first page). The Confidentiality Advisory Group of the Health Research Authority will need to consider any requests for data to be used for purposes other than those specified in our application, so a data request should be accompanied by explanation of the purpose of the request and justification of the public benefit. We also recommend inclusion of a pre-specified plan of analysis.

## Acknowledgements

We thank Katie Ridsdale for clerical assistance with the study, Erica Wallis (Sponsor representative, all members of the Study Steering Committee (Appendix 2) and the site research teams who delivered the data for the study (Appendix 3), and the research team at the University of Sheffield past and present (Appendix 4).

## Role of the funding source

The PRIEST study was funded by the United Kingdom National Institute for Health Research Health Technology Assessment (HTA) programme (project reference 11/46/07). The funder played no role in the study design; in the collection, analysis, and interpretation of data; in the writing of the report; and in the decision to submit the article for publication. The views expressed are those of the authors and not necessarily those of the NHS, the NIHR or the Department of Health and Social Care.

## Appendix 1: scoring triage tools

### Paediatric Observation Priority Score (POPS)

POPS is scored using to the chart below

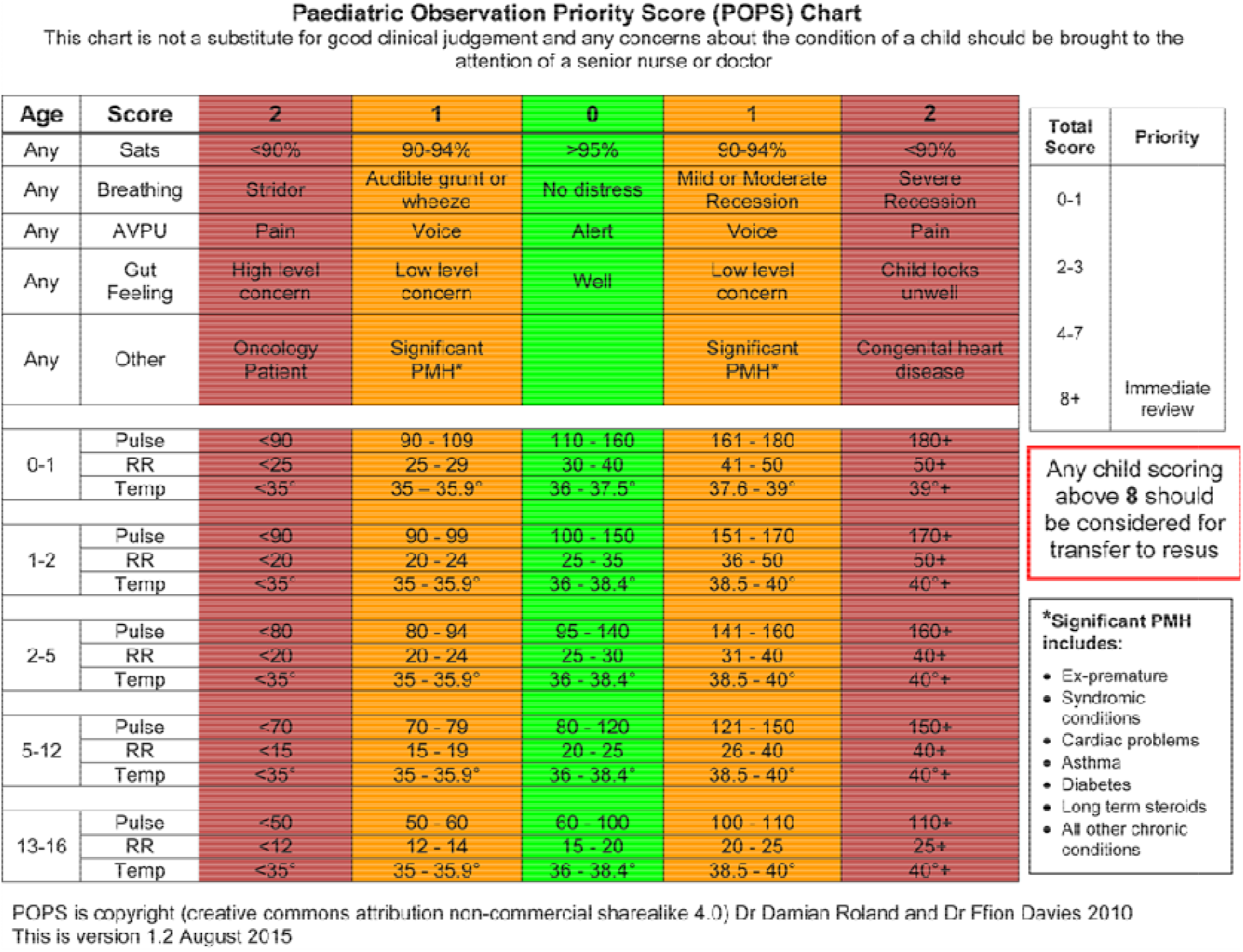

We modified POPS as follows to accommodate limitations of data collection:

- Gut feeling was not scored because its subjectivity made it difficult to reproduce or record from clinical records
- Breathing was scored as two points if either severe respiratory distress or respiratory exhaustion was recorded, otherwise it was scored as zero
- Other was scored as one point if any of prematurity, heart disease, asthma, diabetes, steroid therapy or other chronic lung disease was recorded, and two points if immunosuppression or active malignancy was recorded

If data for a parameter was missing, we assumed it was negative. We excluded the patient from analysis if fewer than three of the parameters were available.

#### Children’s Observation and Severity Tool (COAST)

COAST is scored using age-specific ranges based on separate charts for infants (0-1years), preschool (1-4 years), school age (5-12 years) and teenage (13-18 years), with one point allocated for each of the following:

- Doctor/nurse/family concern
- Abnormal heart rate for age
- Abnormal respiratory rate for age
- Abnormal oxygen saturation
- Moderate or severe respiratory distress
- Altered consciousness
- Pain score

We modified COAST as follows to accommodate limitations of data collection:

- Pain score was excluded
- Only severe respiratory distress was scored
- Only parental concern (parental anxiety) was scored

COAST was therefore scores out of a possible six points. If data for a parameter was missing, we assumed it was negative. We excluded the patient from analysis if fewer than three of the parameters were available.

#### The WHO decision-making algorithm

The WHO decision-making algorithm for hospitalisation with pneumonia recommends admission for children (rule positive) if any of the following are present:

- respiratory rate: >30/minute if over 5 years old; ≥ 40/min if 1-5 years old; or ≥ 50/min if <1year old
- oxygen saturation <90%
- respiratory distress, respiratory exhaustion or severe dehydration recorded
- AVPU is P or U, or GCS<13
- any of the following comorbidities are present; diabetes, cardiovascular disease, chronic respiratory disease, renal impairment, immunosuppression

If data for a parameter was missing, we assumed it was negative. We excluded the patient from analysis if fewer than three of the parameters were available.

#### The Swine Flu Hospital Pathway for Children

The Swine Flu Hospital Pathway for Children consists of seven criteria operating as a rule, with the rule being positive if any criteria reaches its threshold.

| Criteria Label | Criteria | Threshold |
| --- | --- | --- |
| A | Severe respiratory distress | Lower chest wall indrawing, sternal recession, grunting or noise breathing when calm (severe respiratory distress ticked on form) |
| B | Respiratory rate | 50+ breaths per minute if under 1, 40+ breaths per minute if 1+ years |
| C | Oxygen saturation | ≤92% on pulse oximetry, breathing air or on oxygen |
| D | Respiratory exhaustion | Exhaustion or apnoeic episode (20+ second pause in breathing) (respiratory exhaustion ticked on form) |
| E | Dehydration or shock | Sternal capillary refill time >2 seconds, reduced skin turgor, sunken eyes or fontanelle |
| F | Altered conscious level | Strikingly agitated or irritable, seizures or floppy infant |
| G | Other clinical concern | Another clinical concern |

We defined dehydration or shock as being present if severe dehydration was recorded or central capillary refill was categorised as abnormal. We defined altered conscious as GCS is less than 15 or AVPU is anything other than alert.

We were unable to identify other clinical concern from the data, so we evaluated a modified Swine Flu Hospital Pathway using the remaining six criteria.

If data for a parameter was missing, we defined the parameter as being negative. We excluded the patient from analysis if fewer than three of the parameters were complete.

## Notes

### Clinical Trial

ISRCTN28342533

### Clinical Protocols

https://www.sheffield.ac.uk/scharr/research/centres/cure/priest

http://www.isrctn.com/ISRCTN28342533

https://www.fundingawards.nihr.ac.uk/award/11/46/07

### Author Declarations

The North West - Haydock Research Ethics Committee gave a favourable opinion on the PAINTED study on 25 June 2012 (reference 12/NW/0303) and on the updated PRIEST study on 23rd March 2020. The Confidentiality Advisory Group of the Health Research Authority granted approval to collect data without patient consent in line with Section 251 of the National Health Service Act 2006.

### Summary of Updates

Added an author (statistician) that should have been included initially.

